# Training in cortically-blind fields confers patient-specific benefit against retinal thinning after occipital stroke

**DOI:** 10.1101/2023.12.19.23298260

**Authors:** Berkeley K. Fahrenthold, Matthew R. Cavanaugh, Madhura Tamhankar, Byron L. Lam, Steven E. Feldon, Brent A. Johnson, Krystel R. Huxlin

## Abstract

**Purpose:** Damage to the adult primary visual cortex (V1) causes vision loss in the contralateral hemifield, initiating a process of trans-synaptic retrograde degeneration (TRD). Here, we examined retinal correlates of TRD using a new metric to account for global changes in inner retinal thickness, and asked if perceptual training in the intact or blind field impacts its progression.

**Methods:** We performed a meta-analysis of optical coherence tomography (OCT) data in 48 participants with unilateral V1 stroke and homonymous visual defects, who completed clinical trial NCT03350919. After measuring the thickness of the macular ganglion cell and inner plexiform layers (GCL-IPL), and the peripapillary retinal nerve fiber layer (RNFL), we computed individual laterality indices (LI) at baseline and after ∼6 months of daily motion discrimination training in the intact- or blind-field. Increasingly positive LI denoted greater layer thinning in retinal regions affected *versus* unaffected by the cortical damage.

**Results:** Pre-training, the affected GCL-IPL and RNFL were thinner than their unaffected counterparts, generating LI values positively correlated with time since stroke. Participants trained in their intact-field exhibited increased LI_GCL-IPL_. Those trained in their blind-field had no significant change in LI_GCL-IPL_. LI_RNFL_ did not change in either group.

**Conclusions:** Relative shrinkage of the affected *versus* unaffected macular GCL-IPL can be reliably measured at an individual level and increases with time post-V1 stroke. Relative thinning progressed during intact-field training, but appeared to be halted by training within the blind field, suggesting a potentially neuroprotective effect of this simple behavioral intervention.

## Introduction

Cortical blindness (CB) following unilateral damage to the primary visual cortex (V1) or its immediate afferents presents as a homonymous, contra-lesional visual field defect. Although partial recovery can occur spontaneously in the first few months after damage ^1–4^, there are no widely-accepted, validated treatments for the resulting visual defect^5^. Standard of care remains “no intervention”, although occasionally, patients are prescribed compensatory (e.g. saccadic) training or substitution (e.g. prism lenses) therapies ^6–9^. Research also continues to show that visual perceptual training can partially restore vision in CB, measurable by both clinical perimetry and psychophysical tests of visual performance^10–21^.

The importance of developing some form of restorative therapy for CB is further highlighted by burgeoning evidence that once patients reach the chronic stage of >6 months post-stroke, visual field defects do not remain completely stable, as was initially thought ^22^. Instead there appears to be progressive worsening of the perimetrically-defined blind field (BF) without intervention^11,19,22,23^. The most plausible explanation for such deterioration of the BF over time is TRD, which involves the progressive shrinkage and even die-back of neurons in the early visual pathways^24–31^. In humans, structural MRI analyses have shown that the optic tract ipsilateral to occipital cortex damage is often reduced in size ^25,29,30,32–35^, as are the thicknesses of the ganglion cell and nerve fiber layers in corresponding regions of the retina in each eye^24,28–32,34,36–45^.

Retinal ganglion cells (RGCs) are responsible for pre-processing and ferrying visual information to the rest of the visual system. As such, their loss or dysfunction could significantly threaten the potential to recover visual functions in participants with V1 damage. Specifically, retinal neurons in the therapeutically-targetable retino-geniculate-striate pathways susceptible to trans-synaptic retrograde degeneration (TRD) after occipital stroke. Isolating specific consequences of TRD in retinal regions known to synapse with V1-lesion-projecting neurons in the lateral geniculate nucleus is crucial to better understand the relationship between TRD and visual retraining. Approaches to re-train the visual deficit have been shown to confer perimetrically-computed improvements to CB visual fields ^16,46^. However, most literature has focused on benefits to visual perception resulting from visual retraining, with limited knowledge of the effects of training on anatomical substrates of vision ^16,46^. If visual training strengthens existing circuitry or recruits neuronal neighbors, similar to rehabilitation for motor stroke^47–49^, this could potentially impact retinal cells that provide input to residual visual pathways. As such, the present study asked two questions: 1) what is the extent and time-course of relative thinning in affected *versus* unaffected inner retinal layers in humans with unilateral occipital strokes, and 2) does the stimulation afforded by visual training impact the progression of inner retinal thinning in such stroke patients? To answer these questions, we performed a meta-analysis of optical coherence tomography (OCT) data collected as part of a recently-completed, multicenter, randomized, double-masked, clinical trial titled the “Hemianopia Intervention Study” (HIS; ClinicalTrials.gov identifier, NCT03350919). The HIS clinical trial design and results have been published in detail ^50^, but in brief, the trial involved 2 pre-training clinic visits to establish eligibility and measure baseline parameters, a 6-months at-home phase during which training was administered to either the intact field (IF) or BF, and 1 post-training clinic visit to evaluate the effect of training. The primary outcome measure for the HIS clinical trial was change in the 24-2 Humphrey perimetric mean deviation (PMD) from baseline, with significant improvements reported for people trained in their BF, and not those trained in their IF. ^50^ However, the trial also performed OCT imaging and collected measurements of GCL-IPL and RNFL thicknesses in the affected and unaffected retina of each eye in each participant at each time-point. This rich data set provided us a unique opportunity to both measure the extent of TRD in this patient cohort, and analyze the impact of two different of visual training interventions on TRD progression.

## Methods

### Participants

The HIS trial (NCT03350919) recruited 48 CB participants (see **Table 1** for demographics) at 3 US academic medical centers: 20 at the University of Rochester’s Flaum Eye Institute, 18 at the University of Pennsylvania’s Scheie Eye Institute and 8 at the University of Miami’s Bascom Palmer Eye Institute. All procedures were approved by the Western Institutional Review Board (WIRB#1181904), adhered to the tenets of the Declaration of Helsinki and all participants gave written, informed consent.

Participants were between 21 and 75 years of age, with an MRI-confirmed occipital lesion resulting in unilateral homonymous hemianopia. Additionally, their lesions had to have occurred after the age of 18 and a minimum of 90 days prior to screening. Participants were also required to reliably fixate with both eyes during psychophysical testing and clinical Humphrey perimetry, with fixation losses, false-negative, and false-positive rates during perimetry of <20%. Participants were excluded from the study if they presented with any ocular or neurologic disease that would interfere with training. Concurrent use of any other form of visual therapy, or of medications that would affect training were additional exclusion criteria.

### HIS clinical trial design and training intervention

As mentioned earlier, the HIS clinical trial ^50^ involved 2 pre-training clinic visits, a 6-month at-home training phase and 1 post-training clinic visit. While the primary outcome measure was change in the 24-2 Humphrey PMD from baseline to 6 months post-training, OCT data were also collected at each study visit, followed by computerized psychophysical testing focused on instructing participants to correctly perform the training task. Once enrolled, participants were randomized to 2 training arms: intact field (IF) or blind field (BF) training - in a 1:1 ratio, using a permuted block design stratified by site. Participants randomized to these training groups did not differ in age (BF-trained: 56±12 years, range 32-72 years; IF-trained: 61±9 years, range 45-74 years; unpaired t-test p= 0.0990, CI _95_=-11.83 to 1.057) or time since stroke (BF-trained: 41±82 months, range 3-373 months; IF-trained: 43±72 months, range 3-338 months; unpaired t-test p=0.9096, CI_95_=-50.37 to 44.98).

The training intervention was a 2-alternative, forced-choice (2AFC), direction discrimination task using random dot stimuli presented either inside the BF or at a corresponding location in the IF (**Table 1, Supplementary Materials, Supplementary Fig. S1**). During the home training segment, two participants withdrew and their data are not included herein. The two cohorts trained for a comparable number of days (unpaired t-test p=0.3598, CI_95_=-43.82 to 16.27; BF-trained: 101±46 days; IF-trained: 115±51 days).

**Table 1.** Participant demographics. . m: male, f: female, R: right, L: left.

| Subject code | Sex | Age range (years) | Time since stroke (months) | Affected visual hemifield | Training Group |
| --- | --- | --- | --- | --- | --- |
| CB1 | m | 46-50 | 32 | R | Intact |
| CB2 | f | 46-50 | 13 | R | Intact |
| CB3 | m | 70-75 | 20 | L | Intact |
| CB4 | m | 60-65 | 61 | L | Intact |
| CB5 | m | 60-65 | 43 | L | Intact |
| CB6 | m | 55-60 | 105 | R | Intact |
| CB7 | f | 70-75 | 338 | L | Intact |
| CB8 | m | 70-75 | 10 | L | Intact |
| CB9 | m | 66-70 | 6 | R | Intact |
| CB10 | m | 70-75 | 4 | L | Intact |
| CB11 | m | 70-75 | 58 | L | Intact |
| CB12 | f | 66-70 | 16 | R | Intact |
| CB13 | f | 50-55 | 24 | L | Intact |
| CB14 | m | 60-65 | 130 | L | Intact |
| CB15 | m | 56-60 | 13 | L | Intact |
| CB16 | m | 46-50 | 63 | L | Intact |
| CB17 | m | 60-65 | 8 | L | Intact |
| CB18 | f | 46-50 | 15 | R | Intact |
| CB19 | m | 60-65 | 20 | R | Intact |
| CB20 | m | 40-45 | 5 | R | Intact |
| CB21 | m | 66-70 | 3 | R | Intact |
| CB22 | m | 56-60 | 4 | R | Intact |
| CB23 | m | 70-75 | 6 | L | Intact |
| CB24 | m | 66-70 | 60 | L | Blind |
| CB25 | f | 50-55 | 38 | R | Blind |
| CB26 | f | 50-55 | 18 | L | Blind |
| CB27 | m | 40-45 | 15 | R | Blind |
| CB28 | m | 50-55 | 373 | R | Blind |
| CB29 | m | 60-65 | 5 | R | Blind |
| CB30 | m | 70-75 | 47 | R | Blind |
| CB31 | m | 56-60 | 8 | L | Blind |
| CB32 | m | 60-65 | 47 | L | Blind |
| CB33 | m | 40-45 | 11 | R | Blind |
| CB34 | m | 56-60 | 4 | L | Blind |
| CB35 | f | 40-45 | 10 | R | Blind |
| CB36 | m | 66-70 | 105 | L | Blind |
| CB37 | m | 66-70 | 36 | R | Blind |
| CB38 | m | 30-35 | 5 | R | Blind |
| CB39 | f | 46-50 | 5 | L | Blind |
| CB40 | f | 60-65 | 3 | L | Blind |
| CB41 | m | 46-50 | 14 | R | Blind |
| CB42 | m | 66-70 | 3 | L | Blind |
| CB43 | m | 70-75 | 6 | L | Blind |

During pre-training, in-clinic assessment, participants received instructions and underwent baseline testing with this task within their intact and blind hemifields, with fixation enforced binocularly using an Eyelink Duo Mobile eye-tracker (SR Research, Mississauga, Ontario, Canada). Training locations were selected at sites where performance first dropped to chance (50% correct) after a 1 ° lateral shift along the x-axis from the intact into the BF. Using the location where performance first drops to chance as a starting point affords proximity to intact circuitry, enhancing the possibility that training may recruit perilesional V1 ^51^, and/or induce plasticity and re-integration of residual, damaged circuitry. IF-training locations were selected to be mirror-symmetric to those chosen for training in the BF. CB participants were then sent home to train and were asked to perform 300 trials of the 2AFC task once daily for a minimum of 5 days per week, at their assigned, training location. Participants trained at a single location at a time, either in the IF or BF. Once performance improved sufficiently (at least 10 sessions at a threshold <25° with a standard deviation of less than 5°), the location was moved 1° laterally away from the vertical meridian. Participant performance as a result of these interventions has been published^50^ and will not be repeated here in detail.

### Humphrey visual field testing and analysis

Each participant’s visual deficit was quantified through Humphrey visual field (HVF) perimetry, which was performed twice in both eyes during each study visit. The University of Rochester and the University of Pennsylvania used a Humphrey Field Analyzer II-i, and the University of Miami used a Humphrey Field Analyzer 3 (Zeiss Humphrey Systems, Atlanta, GA), with all sites using a 24-2 testing pattern. A white, size III stimulus was presented on a background with a luminance of 11.3 cd/m^2^ and thresholds were calculated with the Swedish Interactive Threshold Algorithm (SITA-standard). Participants’ visual acuity was corrected to 20/20 for testing, and fixation was controlled using the gaze/blind spot automatic settings. The first test was excluded in both eyes to account for potential learning effects. If the second field set was not deemed reliable or could not be completed, the first set was used instead. Participants who did not have complete, reliable pre- and post-training visual fields were excluded from the present HVF analyses(n=5); an additional two participants failed to complete training and were also removed from our analysis. Two metrics were derived from HVF tests: the perimetric mean deviation (MD) and the average luminance detection sensitivity across the entire blind hemifield of vision. The MD is calculated by the perimeter using an internal, weighted variance from age-defined normal population values, to estimate the amount of vision lost across the measured visual field. In the present study, sensitivity thresholds from the blind hemifield (ST_BF_) were additionally averaged in each eye to capture deficit-specific changes. We then took the monocular MD and ST _BF_ and averaged them to generate a binocular (OU) version of each metric, for pre- and post-training comparisons, in order to compare with binocularly-computed OCT laterality indices.

### Optical coherence tomography (OCT) procedures and analysis

Retinal OCT was performed using Cirrus HD machines (Carl Zeiss Meditech) at each study site before and after training. A 512 × 128 Mac Cube scan was used to examine the ganglion cell and inner plexiform layers (GCL-IPL) around the fovea, and 200 × 200 optic nerve cube scans were used to examine the retinal nerve fiber layer (RNFL). Scans were excluded if they failed to meet signal strength ≥7 in each eye.

OCT analyses performed as part of the HIS clinical trial ^50^ differed from those performed here in several ways. First, they involved group-level comparisons of GCL-IPL and RNFL thickness changes pre- to post-training across affected and unaffected retinal regions, separated by blind field sector and for each eye independently. Here, for the GCL-IPL, we combined the 2 nasal sectors together, and the two temporal sectors together. Furthermore, after reviewing OCT raw data, we excluded 3 HIS participants due to retinal folding or epiretinal membrane/RNFL detachment severe enough to impact layer thickness measurements. In remaining participants, we then computed a laterality index (LI) to account for individual, baseline thickness variances using the following formula:

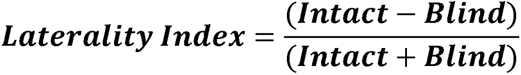

LI was computed for the GCL-IPL using the 2 nasal and 2 temporal sector values of both eyes, excluding superior and inferior sectors that overlapped the vertical meridian (**Fig. 1A**). The nasal and temporal macular segments of each eye corresponding to the blind or intact hemifield were then averaged together according to each participant-specific deficit. For example, a right-sided visual deficit (left-sided occipital lesion) is represented in the nasal sectors of the right eye and the temporal sectors of the left eye (see example in**Fig. 1A**).

**Figure 1.**
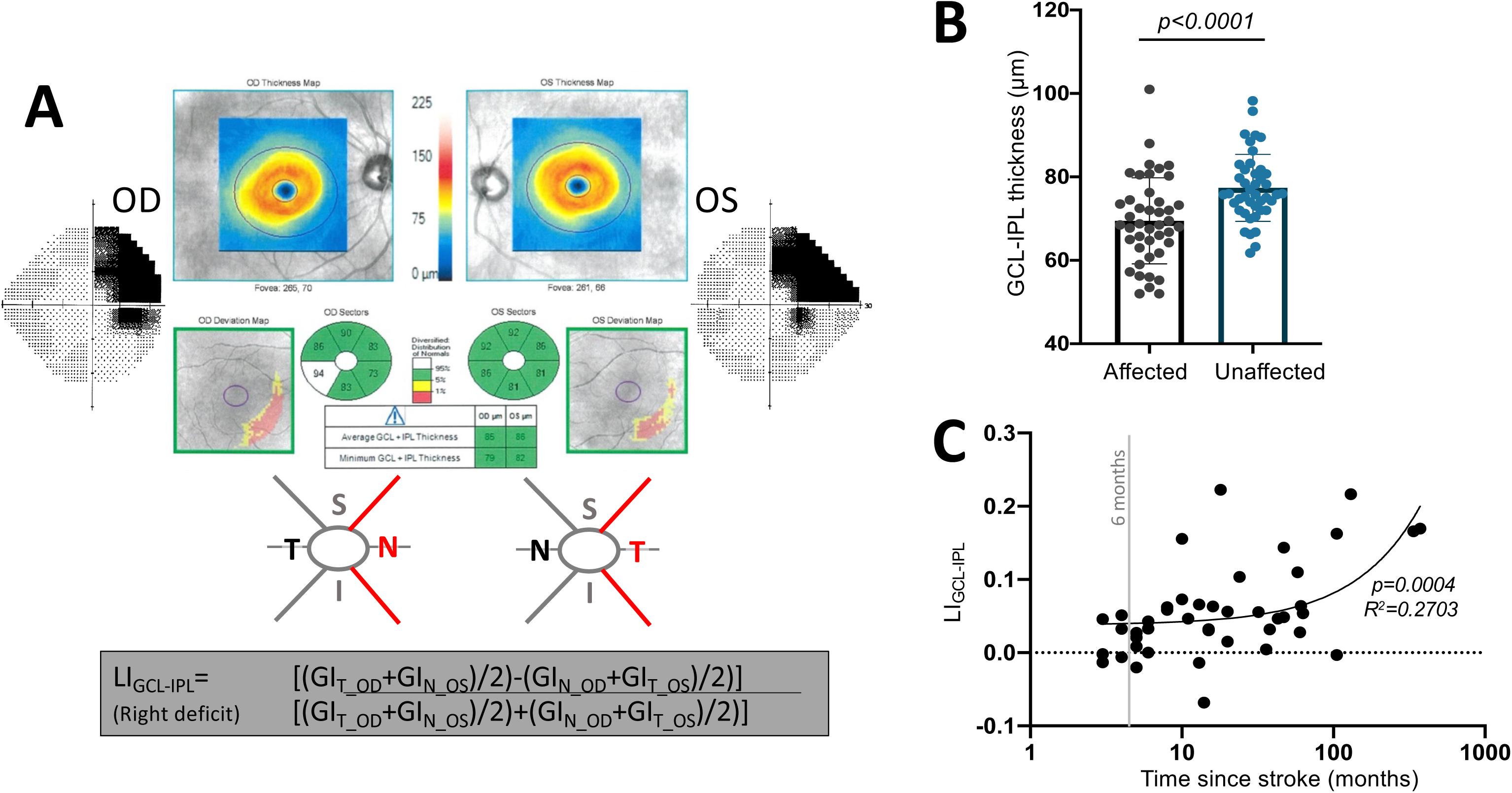
**A:** Computation of LI for the GCL-IPL using the nasal (N) and temporal (T) sector GCL-IPL (GI) thickness values of both eyes, excluding superior (S) and inferior (I) sectors since they overlapped the vertical meridian. **B:** Plot comparing GCL-IPL thicknesses in the affected or unaffected hemiretinas (*paired t-test, CI*_95_ *=5.212 to 10.60, t*_42_ *=5.921, p = <0.0001*). **C:** Plot of LI_GCL-IPL_ against time since stroke (*linear regression, R*^2^*=0.2703, CI (y-intercept)=0.018 to 0.057, p=0.0004)*.

Computing a laterality for the RNFL regions impacted by the deficit attempted to account for the crossed and uncrossed fibers in corresponding peripapillary sections^52^ (**Fig. 2A**). Superior and inferior peripapillary regions comprised of uncrossed fibers, and nasal peripapillary regions comprised of crossed fibers represent intact or blind hemifields. For example, the same right-sided visual deficit area described above was represented by the superior and inferior RNFL sectors of the left eye as well as the nasal RNFL sector of the right eye (**Fig. 2A**).

**Figure 2.**
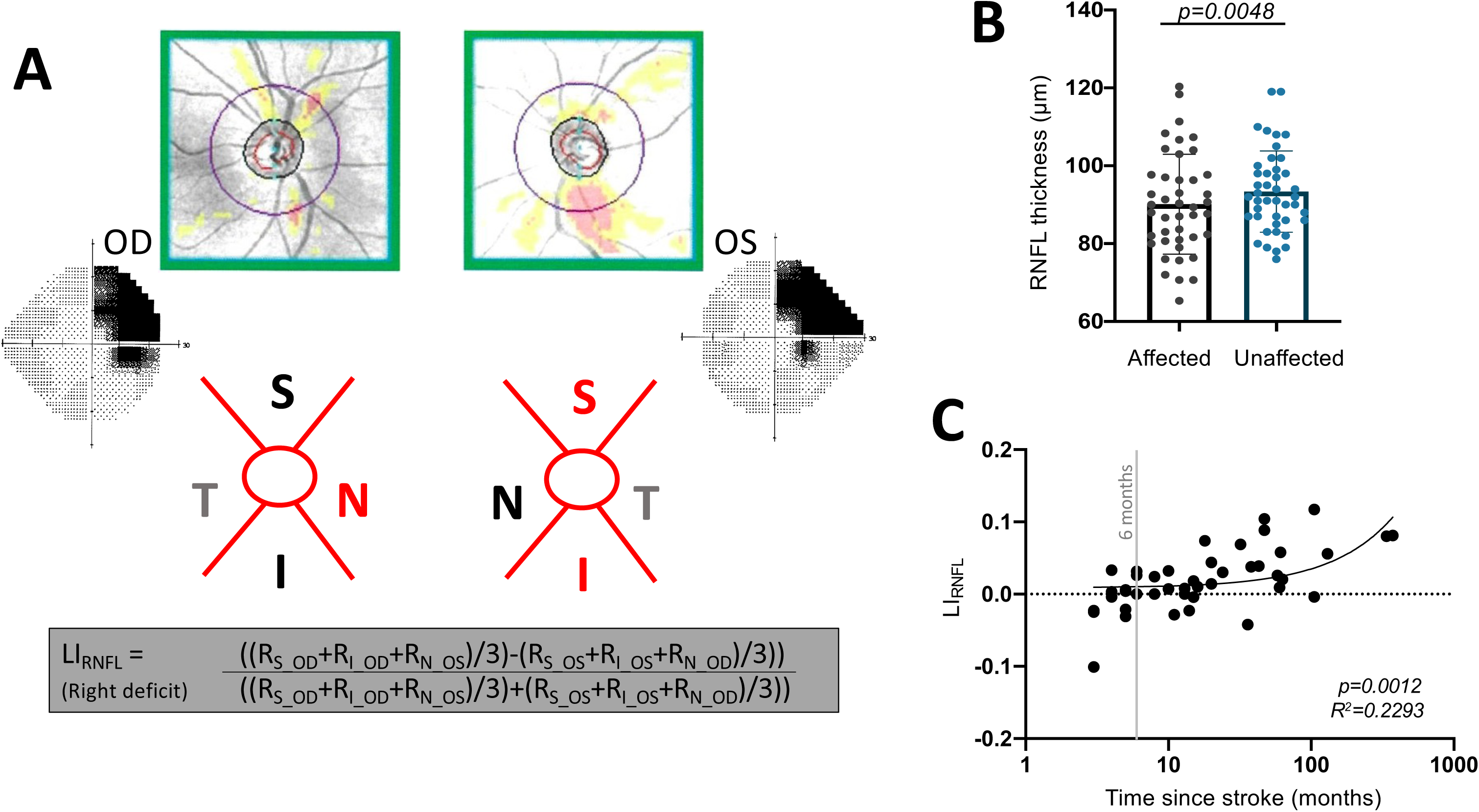
**A:** Computation of LI using RNFL thickness values (R) from superior (S) and inferior (I) peripapillary regions comprised of uncrossed fibers, and nasal (N) peripapillary regions comprised of crossed fibers representing intact or blind hemifields.**B:** Plot comparing affected peripapillary RNFL segments carrying RGC axons representing the visual field defect to unaffected carrying predominantly intact field fibers (*paired t-test, CI*_95_ *=1.046 to 5.450, t*_42_ *= 2.977, p = 0.0048*). **C:** Plot of LI_RNFL_ against time since stroke (*linear regression, R*^2^*=0.2293, CI =-0.0004 to 0.0217, p=0.0012)*.

### Statistical Analyses

Paired t-tests were used to assess within-subject differences. For independent sample comparisons, unpaired t-tests were used when contrasting 2 groups. If standard deviations were not the same in each group, Welch’s correction was used. Linear regressions were used to model the relationship between explanatory variables and dependent outcomes, with r values and 95% confidence intervals (CI_95_) for rho provided, and significance estimated using a t-test.

## Results

### Baseline retinal layer thicknesses – effects of time since stroke

Prior to intervention, GCL-IPL thicknesses corresponding to the blind or intact hemifields were significantly different from each other, with the affected hemiretina’s GCL-IPL being thinner than the unaffected hemiretina’s (**Fig.1B**). We then computed LI to factor out possible global retinal phenomena (e.g., aging-related, metabolic, etc.) in order to better isolate lesion-specific degeneration in retinal regions corresponding to perimetrically-defined visual deficits. The LI_GCL-IPL_ was positive, averaging 0.056 ± 0.06, with a range of -0.068 to 0.29. An LI of 0 would indicate no relative thinning of the lesion-projecting compared to the non-lesion projecting part of the retina, while positive LI values denote thinning in retinal areas representing the blind hemifield relative to those representing the intact hemifield. Importantly, the LI _GCL-IPL_ was positively correlated with time since stroke (**Fig. 1C**), with greater thinning of the affected hemiretina GCL-IPL in participants imaged beyond 12 months post-stroke compared to those imaged prior to this timepoint (**Fig. S2A**).

A similar pattern of results was obtained for the peripapillary RNFL, which was thinner for segments carrying RGC axons representing the visual field defect compared to those carrying predominantly intact field fibers (**Fig.2B**). As a result, LI _RNFL_ averaged 0.019 ±0.04, ranging from -0.10 to 0.11. Moreover, just like LI_GCL-IPL_, LI_RNFL_ was positively correlated with time since stroke (**Fig. 2C**), with relative thinning most pronounced beyond 12 months post-lesion (**Fig. S2B**). Overall, these data show clear GCL-IPL and RNFL thinning in regions of the retina carrying either RGC somata, dendrites and/or axons representing blind regions of the visual field. They also show greater thinning at later than earlier timepoints, especially >12 months after occipital stroke.

### Effect of visual training on GC complex thickness

We next asked whether visual training altered signs of TRD at the level of the retina. Here, we analyzed CB patients who completed 6 months of visual training as part of the HIS clinical trial^50^. As previously reported, global direction discrimination training in the perimetrically-defined BF of CB patients elicits improvements not only on the trained task, but also on binocular (OU) Humphrey perimetry ^11,50^. Consistent with this observation, participants trained in their BF exhibited a systematic improvement in OU MD (**Fig.3A**). To ascertain if the change in MD was driven by the blind hemifield (*versus* improved ability to perform Humphrey perimetry across the entire test area), we also computed OU ST _BF_ change for the blind hemifield. OU ST _BF_ improved significantly following BF training (**Fig. 3B**), contrasting with a lack of significant changes - for both OU MD and ST _BF_ - in the IF-trained cohort (**Fig. 3D, E**). Notably, a strong correlation exists between MD and ST_BF_ in both cohorts, pre- and post-training (**Fig. 3C, F**).

**Figure 3.**
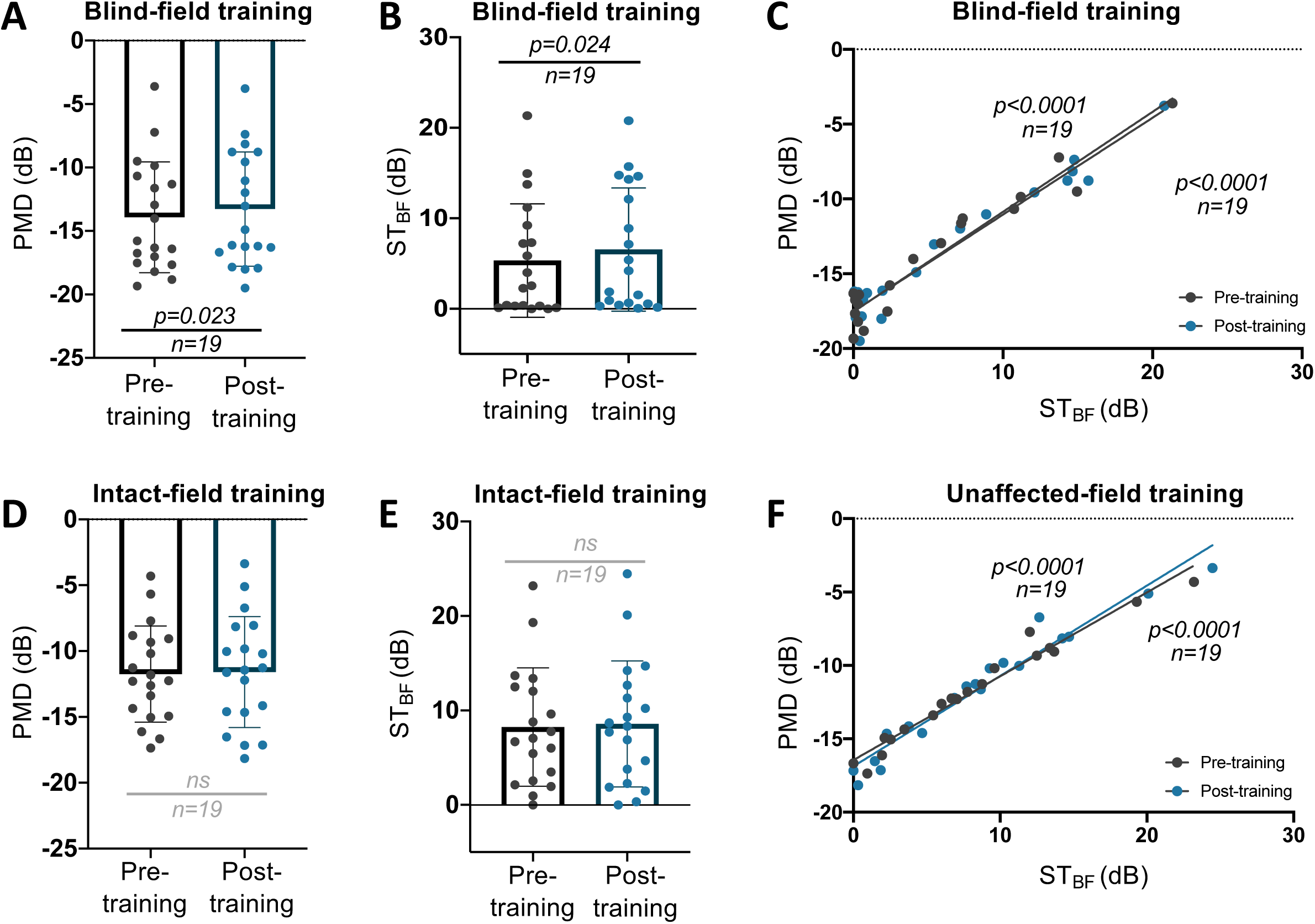
**A:** Plot of pre- and post-training OU MD in BF-trained participants (*paired t-test*, *CI*_95_ *=0.098 to 1.2, t*_18_ *= 2.473, p = 0.023, mean of differences = 0.65*±*1.15)*. **B:** OU ST_BF_ pre- and post- training following BF training *(paired t-test, CI*_95_ *=0.177 to 2.258, t*_18_ *= 2.458, p = 0.024, mean of differences = 1.22*±*2.16).* **C:** Linear regression of MD against ST_BF_ pre-training: *R*^2^*=0.9365, CI (y-intercept)=-18.28 to -16.82;* post-BF training: *p <0.0001; R*^2^*=0.9526, CI (y-intercept)=-18.21 to - 16.84, p <0.0001*. **D:** Plot of pre- and post-training OU MD in IF-trained participants (*paired t-test, CI*_95_ *=-0.3284 to 0.6284, t*_18_ *= 0.6587, p = 0.5184, mean of differences = 1.5*±*0.9926).* **E:** OU ST_BF_ pre- and post-training following IF training(*paired t-test, CI*_95_ *=-0.1917 to 0.8655, t*_18_ *= 1.339, p = 0.1972, mean of differences = 0.3369*±*1.097*). **F:** Linear regression of MD against ST_BF_ pre-training: *R*^2^*=0.963, CI (y-intercept)=-17.04 to -15.87; post-IF training: p <0.0001; R*^2^*=0.9481, CI*_95_*(y-intercept)=-17.67 to -16.08, p <0.0001*). ns: not statistically significant.

Having established a subtle but differential effect of training on perimetry between the two cohorts, we then asked if - and to what degree - the two types of intervention impacted retinal thinning. For LI_GCL-IPL_, there was a significant overall increase pre- to post-training across all participants (**Fig. 4A**). However, no detectable changes occurred pre- to post-training overall in LI_RNFL_ (**Fig. 4B**). Separating the two interventions, LI_GCL-IPL_ was significantly larger post-training in those who trained in their intact field (**Fig.5A**), but not in BF-trained participants (**Fig.5B**). Furthermore, in the IF training group, raw GCL-IPL thicknesses were significantly lower in the post-training affected hemiretina (**Fig. 5C**). Post-training affected GCL-IPL thicknesses also significantly changed from pre-training in the BF training group (**Fig. 5D**). Additionally, a significant difference in magnitude of change in the affected relative to unaffected hemiretinas was present in IF trained participants and, critically, no such difference was found in BF trained participants (**Fig. 5E**). However, we fail to reject the null hypothesis that the pre-post differences of the affected hemiretinas differ by training type (**Fig. 5E**).

**Figure 4.**
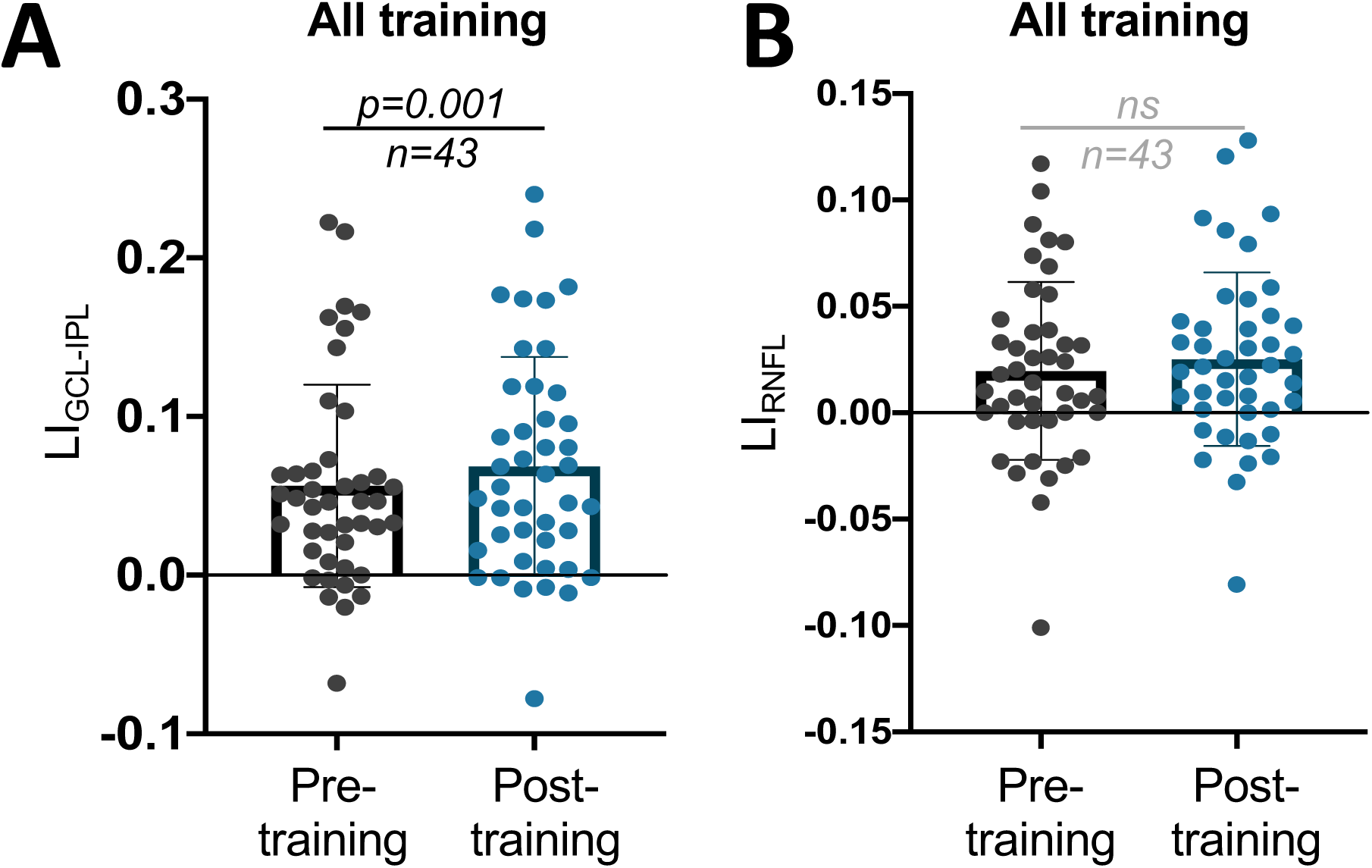
**A:** Plot of LI_GCL-IPL_ pre- to post-training for all participants (*paired t-test, CI*_95_ *=0.005 to 0.019, t*_42_ *= 3.424, p = 0.001*). **B:** Plot of LI_RNFL_ pre- to post-training of all participants (*paired t-test, CI*_95_ *=-0.0015 to 0.01251, t*_42_ *= 1.572, p = 0.1234*). ns: not statistically significant.

**Figure 5.**
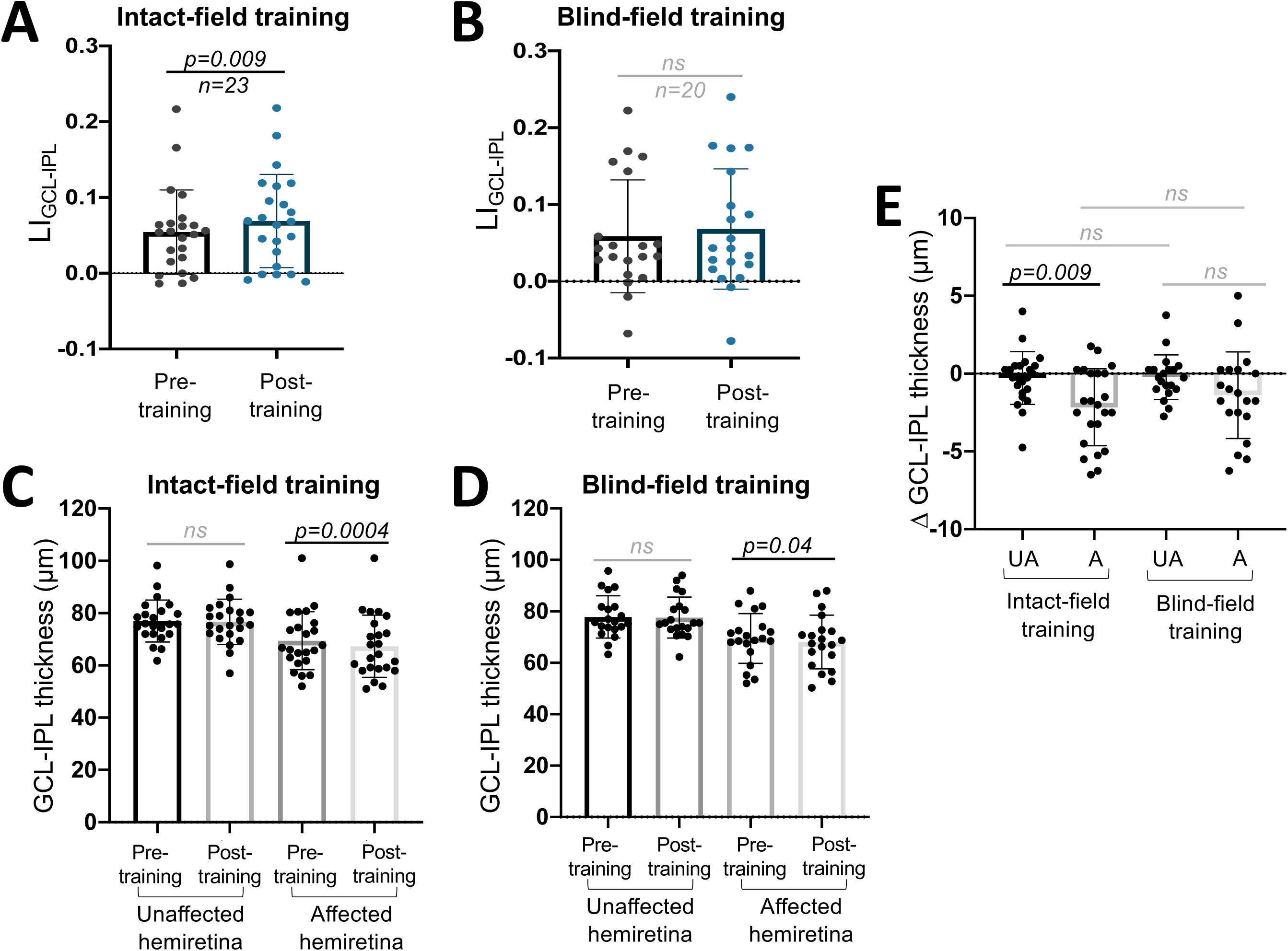
**A:** Plot of LI_GCL-IPL_ pre- to post-training of participants who trained in their IF (*paired t-test, CI*_95_ *=0.003 to 0.025, t*_22_ *= 2.837, p = 0.0096*). **B:** Plot of LI_GCL-IPL_ pre- to post-training of participants who trained in their BF (*paired t-test, CI*_95_ *= -0.0008 to 0.0199, t*_19_ *= 1.916, p = 0.07*). **C:** Comparisons of unaffected and affected hemiretina GCL-IPL thicknesses, before and after training in the IF (*unaffected pre- vs post-training: paired t-test*, *CI*_95_*= -1.017 to 0.4516, p = 0.4332; affected pre- vs post-training: paired t-test, CI*_95_ *= -3.233 to -1.093, p = 0.0004).* **D:** Comparison of unaffected and affected hemiretina GCL-IPL thicknesses, before and after BF training (*unaffected pre- vs post-training: paired t-test, CI*_95_ *= -0.8943 to 0.4443, p = 0.4902; affected pre- vs post-training: paired t-test, CI*_95_ *= -2.691 to -0.0843, p = 0.04).* **E:** Comparison of change in GCL-IPL thickness from pre- to post-training in IF or BF-trained participants (*unaffected vs affected hemiretina in IF-trained subjects, paired t-test, CI*_95_*=0.2063 to 3.555, p=0.009*; *unaffected vs affected hemiretina in BF-trained subjects, CI*_95_*=-2.601 to 0.2762, p=0.1071*; *unaffected vs unaffected of both training groups, unpaired t-test, CI*_95_*=-0.9176 to 1.033, p=0.9056*; *affected vs affected of both training groups, unpaired t-test, CI*_95_*=-0.8438 to 2.395, p=0.3391*). A: Affected, UA: Unaffected, ns: not statistically significant.

When assessing the impact of training on the RNFL, no significant change in LI _RNFL_ was found in either group pre- to post-training (**Fig. 6A, B**). Similarly, no significant pre-post training differences were observed in either training cohort for raw RNFL thickness (**Fig. 6C, D**). Additionally, when assessing pre-post change, no significant differences were seen between or within training groups (**Fig. 6E**).

**Figure 6.**
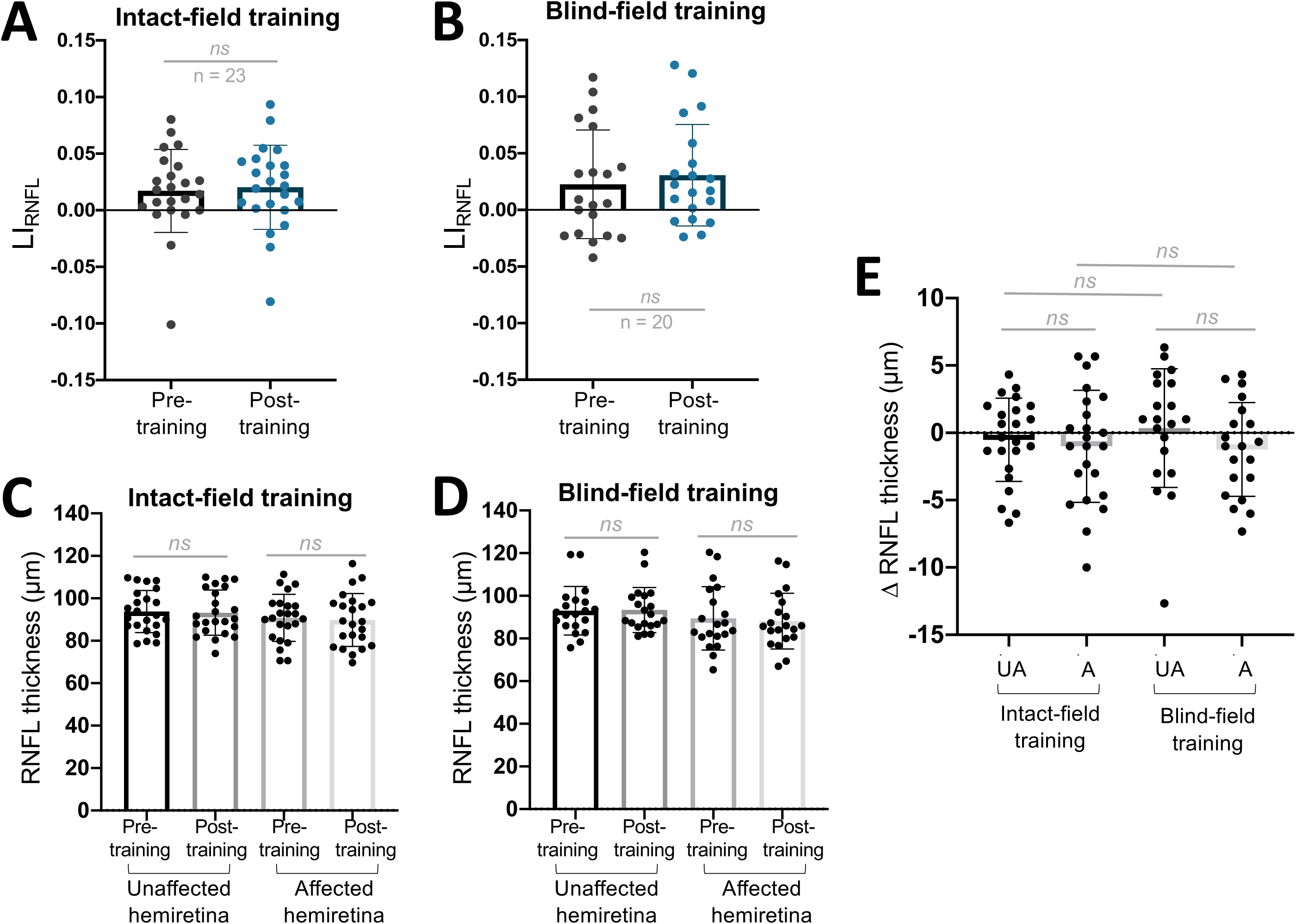
**A:** Plot of LI_RNFL_ pre- to post-training of participants who trained in their IF (*paired t-test, CI*_95_ *=-0.003 to 0.0123, t*_22_ *= 0.7322, p = 0.4718*). **B:** Plot of LI_RNFL_ pre- to post-training of participants who trained in their BF (*paired t-test, CI*_95_ *= -0.0036 to 0.0197, t*_19_ *= 1.446, p = 0.1645*). **C:** Comparison of unaffected and affected RNFL thicknesses, before and after IF training (*unaffected pre- vs post-training: paired t-test, CI*_95_ *= -1.860 to 0.8163, p = 0.4274; affected pre- vs post-training: paired t-test, CI*_95_ *= -2.796 to 0.7962, p = 0.2606).* **D:** Comparison of unaffected and affected RNFL thicknesses, before and after BF training (*unaffected pre- vs post-training: paired t-test, CI*_95_ *= -1.714 to 2.414, p = 0.7266; affected pre- vs post-training: paired t-test, CI*_95_ *= -2.862 to 0.3951, p = 0.1294).* **E:** Comparison of change in RNFL thickness from pre- to post-training in IF or BF-trained participants (*unaffected vs affected in IF-trained subjects: paired t-test, CI*_95_*=-2.096 to 1.139, p=0.546*; *unaffected vs affected in BF-trained subjects: paired t-test, CI*_95_*=-3.627 to 0.4601, p=0.1213*; *unaffected vs unaffected of both training groups: unpaired t-test, CI*_95_*=-1.451 to 3.195, p=0.4529*; *affected vs affected of both training groups: unpaired t-test, CI*_95_*=-2.614 to 2.148, p=0.8441*). A: Affected, UA: Unaffected, ns: not statistically significant.

Consistent with these findings, changes in ST _BF_ and LI _GCL-IPL_ were directly [and inversely] correlated in those trained in their blind hemifield (**Fig. 7A**), but not in those trained in their IF (**Fig. 7B**). No significant correlations were observed between changes in ST_BF_ and LI_RNFL_ in either training cohort (**Fig. 7C, D**).

**Figure 7.**
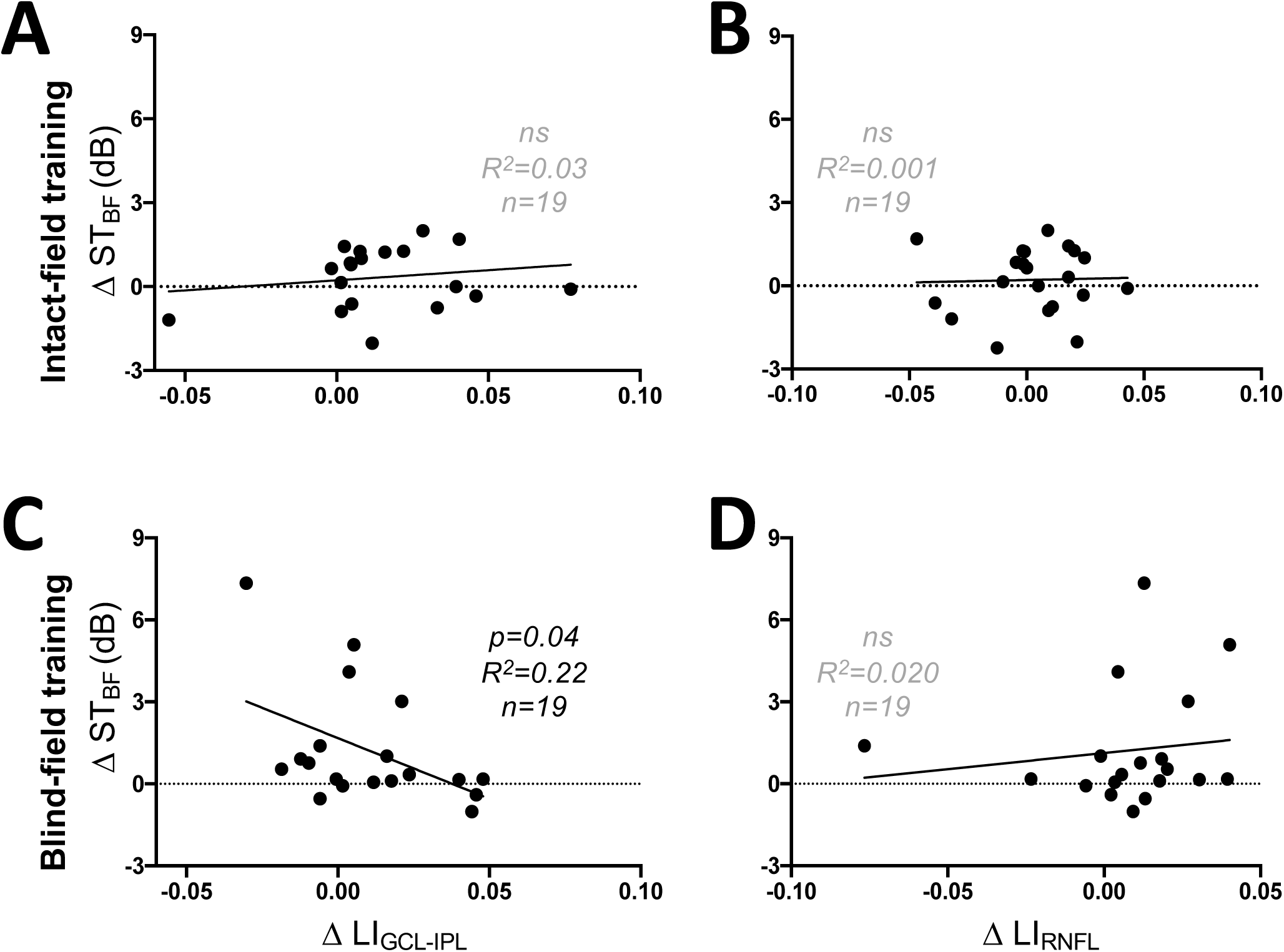
**A:** Plot of change in ST_BF_ against LI_GCL-IPL_ of participants trained in their IF (*linear regression, R*^2^*=0.0307, CI*_95_ *(y-intercept)=-0.3997 to 0.8511, p=0.4727).* **B:** Plot of changes in ST_BF_ and LI_RNFL_ in IF-trained participants (*linear regression, R*^2^*=0.001, CI (y-intercept)=-0.3497 to 0.766, p=0.8857*). **C:** Plot of change in ST_BF_ against LI_GCL-IPL_ of participants trained in their BF (*linear regression, R*^2^*=0.2170, CI (y-intercept)=0.6239 to 2.723, p=0.04)*. **D:** Plot of changes in ST_BF_ and LI_RNFL_ in participants training in the BF(*linear regression, R*^2^*=0.0196, CI (y-intercept)=0.0093 to 2.241, p=0.5674*).

## Discussion

The present study asked – for the first time – whether visual stimulation provided by perceptual training alters the progression of retinal ganglion cell layer complex thinning after str oke damage to the occipital cortex in adult humans. First, we confirmed prior reports of relative thinning in the affected *versus* unaffected retinas’ GCL-IPL and RNFL after unilateral V1 damage^28,29,31,32,34,36,39,41,53^ using non-invasive OCT imaging. Second, the spread of post-stroke times at participant enrollment allowed us to define a time-course for this thinning. Finally, we now provide evidence that a simple behavioral intervention slows or blocks the progression of relative GCL-IPL thinning whereas comparable stimulation of the intact hemifield of vision fails to do so.

### Occipital damage causes variable, progressive shrinkage of the GC complex

Our observations showed that the largest, positive deviations from 0 in LI _GCL-IPL_ and LI _RNFL_ occurred beyond 12 months post-stroke. While some deviation in LI _GCL-IPL_ (but not LI _RNFL_) was also observed in our earliest participants, there was considerable inter-individual variability which precluded a significance analysis in the present cohort. Large deviations of LI values from 0 were previously observed for optic tract volumes using structural magnetic resonance imaging, starting from ∼6 months post stroke, albeit also with large inter-individual variability^35^. This time-course differential makes some sense if one considers that the optic tract contains the distal portions of RGC axons, right before they synapse in the dorsal lateral geniculate nucleus. These distal axons, being closer to the V1 lesion site, might exhibit earlier signs of target loss and degeneration than the cell bodies and dendritic arbors of the parent cells in the retina, but once again, a larger sample size earlier post-stroke would be needed to make this determination from a statistically-valid standpoint.

As stated earlier, a positive LI reflects a relative thinning of the lesion-projecting *vs.* intact-hemisphere-projecting portions of the macular GCL-IPL. This relative thinning could be attributable to shrinking of the RGC soma, cell death, and/or changes in cell branching; similarly, relative thinning in the RNFL could result from RGC axonal loss or shrinkage or both^24,28,29,32,34,37,40,53–55^. Though past studies show that RGCs are ultimately lost over time after occipital damage^28,29,31,55,56^, there is also evidence that RGCs change size based on metabolic activity or the beginning stages of apoptosis ^57,58^. As such, slowing or even reversing retinal thinning may be possible if intervention occurs prior to significant cell death.

Importantly, both the GCL-IPL and RNFL were previously reported to thin with increasing age in humans^59^, a fact confirmed in the present data set, and is likely related to cell loss and/or shrinkage (**Fig. S3**). However, it is important to note that by computing and tracking changes in LI rather than raw layer thicknesses, we were able to dissociate the impact of the occipital stroke and subsequent training interventions, from this natural trend.

### Visual training blocks the progression of *relative* GC complex thinning

In spite of initial retinal ganglion cell complex thinning at baseline, participants who trained in their BF for 6 months showed improvements in binocular performance metrics derived from Humphrey perimetry and seemed to avoid the increase in LI_GCL-IPL_ that occurred in participants randomized to train in their IF. While GCL-IPL thickness decreased in both groups, the change in GCL-IPL thickness of the affected relative to the unaffected hemiretina was only significant in the IF-trained participants. Coupled with a failure to reject the null hypothesis in pre- to post-training differences of LI_GCL-IPL_ in the BF-trained group, this suggests a subtle but significant effect of training location on GCL-IPL thinning within a given patient, which is lost when comparing effects across individuals. The inherent variability in OCT layer thickness in small cohorts makes it difficult to compare groups directly. This is further complicated by variability introduced due to time-dependent TRD. In the future, increasing the sample size to increase sensitivity is crucial to better understanding the anatomical underpinnings of visual retraining. This is a difficult endeavor with two critical limitations 1) CB participants with lesions limited to the occipital cortex are rare and challenging to recruit and 2) once recruited, CB participants require time-intensive testing and evaluation. Alleviating these limitations would require expansion of collaborating facilities and personnel, as well as relaxing inclusion and exclusion recruitment criteria, leading to a more heterogenous patient population. However, despite current limitations these within-group comparisons provide novel insights into training-dependent changes within the early visual pathway.

These surprising observations suggest first that OCT imaging and our derived LI metric is a sensitive biomarker for assessing the impact of training in post-stroke CB patients. Just as importantly, it also suggests that an intervention which locally stimulates RGCs in a retinal area deprived of several key central targets may benefit the structural integrity of these residual cells. In turn, this may increase the likelihood that these neurons are retained long-term in the residual visual circuitry, perhaps providing the neural substrates of training-induced recovery of visual functions seen deeper into the visual deficit^51^. Conversely, training within the intact field locally stimulates circuits that are not directly affected by V1 damage-mediated TRD^26,29,43^. Though V1 areas of both hemispheres representing visual information along the vertical meridian are connected via callosal axonal projections^60^, notable due to the training locations of these participants, these interhemispheric connections do not appear to provide enough benefit to anterior portion of the visual pathway to be observable at the level of the retina.

So, what could underlie the stabilization of LI _GCL-IPL_ in BF-trained participants? As mentioned earlier, damaged RGCs undergo changes in their dendritic arbors in the IPL ^61,62^ and in supporting cells, such as muller glia ^57,58,63^, which span the entire thickness of the retina. Training in the BF could increase the energy demands of stimulated RGCs, and by consequence, of surrounding supporting cells, in turn causing structural changes manifested as a cell-size increase and/or shrinkage prevention ^36,57^. Changes in surviving RGCs are of course likely occurring in tandem with RGC loss due to retrograde degeneration – a phenomenon on which visual training’s effects are unknown.

An important question emerging from the present results is whether the stabilization of the LI _GCL-IPL_ persists after BF training stops. If this phenomenon relies on increased retinal activity due to training, it is possible that physiological mechanisms of TRD will eventually overcome the benefits gained once training ceases. However, it is also possible that if participants incorporate their regained visual abilities into everyday usage, they could maintain them and sustain their associated circuits.

Finally, we saw no significant changes in LI _RNFL_ or RNFL thickness in either training cohort, although several factors likely limited our ability to detect such changes with OCT, including the anatomical complexities of the RNFL in different peripapillary zones, the very small volume of the RNFL overall, our relatively small sample size and inter-subject variability. Future studies using larger sample sizes, more detailed analyses and better imaging resolution will be required to rigorously elucidate the impact of training on the RNFL.

## Conclusion

In conclusion, the present work investigated the impact of a visual training intervention administered either inside the blind or intact field of occipital stroke patients on the progression of TRD at the level of the inner retina. We found that relative thinning in the GCL-IPL and RNFL mirrored a distinct time-course post-stroke previously reported in the literature. Training for ∼6 months with a motion discrimination task inside the blind hemifield appeared to block the progression of relative thinning in the ganglion cell complex. In contrast, this relative thinning proceeded unabated when training was administered to the intact field of vision. Our results provide the first evidence of a greater structural benefit in the retina for a behavioral intervention that stimulates circuitry impacted by V1 damage, over one that stimulates the intact circuitry.

## Data Availability

All data produced in the present study are available upon reasonable request to the authors

## Non-standard Abbreviations and Acronyms

CB: Cortical Blindness
OCT: Optical Coherence Tomography
LI: Laterality Index
dLGN: dorsal lateral geniculate nucleus
GCL-IPL: Ganglion Cell Layer-Inner Plexiform Layer
RGC: Retinal Ganglion Cell
HIS: Hemianopia Intervention Study
HVF: Humphrey Visual Field
OU: Oculus Uterque (both eyes)
OS: Oculus Sinister (left eye)
OD: Oculus Dextrus (right eye)
SEM: Standard Error Mean
PMD: Perimetric Mean Deviation
ST_BF_: Mean Sensitivity of Blind Field
TRD: Trans-synaptic Retrograde Degeneration

