## Supplemental Materials for "Training in cortically-blind fields confers patient-specific benefit against retinal thinning after occipital stroke"

**Supplementary Materials**

**Supplemental Methods**

**HIS Trial Task (Supplementary Figure 1)**

Participants were asked to discriminate the upward or downward deviation from horizontal of a patch of moving dots, irrespective of the base left or right directional component of their global motion. Stimuli consisted of 100% coherently-moving black dots on a mid-grey background, within a 5° diameter circular aperture, at a density of 3 dots/deg^2^. Dots moved at a speed of 10deg/sec, with a 250ms lifetime, over the stimulus duration of 500ms. A tone indicated stimulus onset. Participants responded using arrows on a keyboard, denoting their perceived direction of motion (up or down). Auditory feedback was provided on every trial to indicate correct and incorrect responses.

Task difficulty was adjusted using a 3:1 staircase that decreased the size of the angle between the direction of motion and the horizontal meridian as follows: 90°, 75°, 60°, 45°, 30°, 25°, 15°, 10°, 5°, 2.5°, 1°. Each trial began with fixation of a centrally-presented target for 1000ms. A white triangle (feature-based attention pre-cue) was presented 500ms before stimulus onset for 200ms, either to the left or right of the fixation point, indicating the base direction of motion (left or right). The height of the triangle changed to reflect the approximate difficulty of the upcoming trial, with a large cue representing a larger deviation from horizontal (i.e., an easier trial), and a small cue representing a smaller upcoming angle of discrimination (i.e., a harder trial).

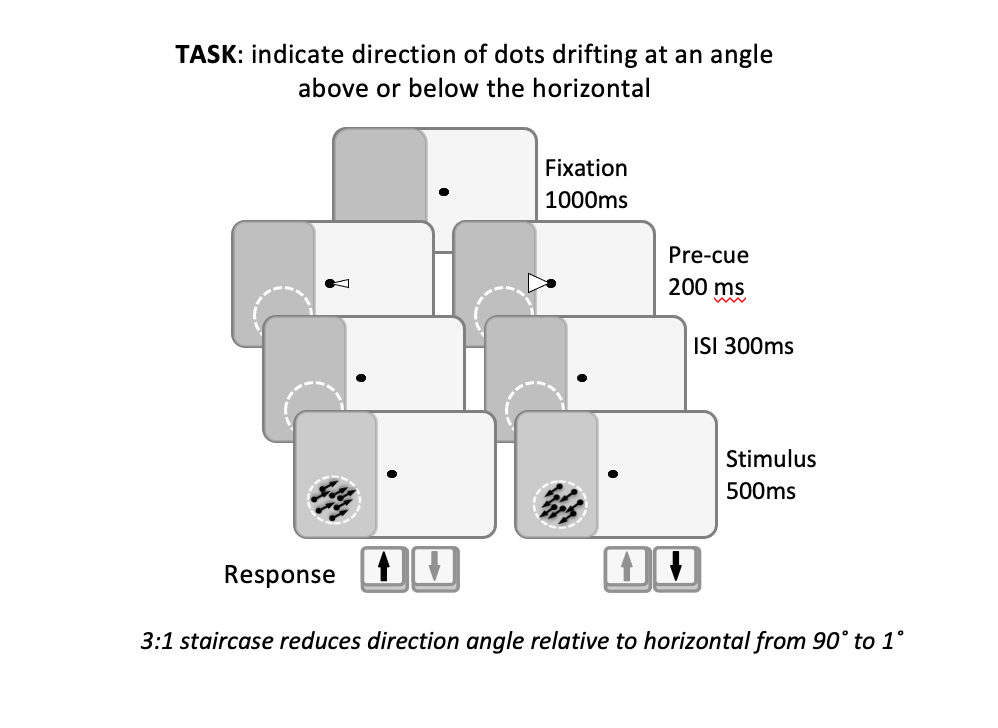

**Supplementary Figure 1.** Schematic of motion discrimination dot task used as a training intervention.

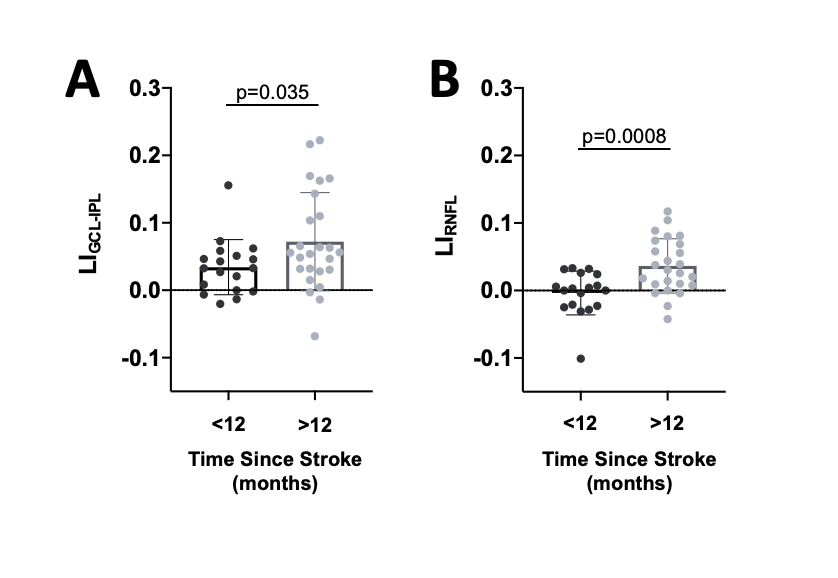

**Supplementary Figure 2.** **A:** Plot comparing pre-training LI_GCL-IPL_ values of participants less than and greater than 12 months post-stroke (*Welch’s unpaired t test: CI_95_ =0.0026 to 0.0732, t_39.08_ = 32.174, p = 0.0358*). **B:** Plot comparing pre-training LI_RNFL_ values of participants less than and greater than 12 months post-stroke (*Welch’s unpaired t test: CI_95_ =0.0176 to 0.0624, t_40.43_ = 3.614, p = 0.0008).*

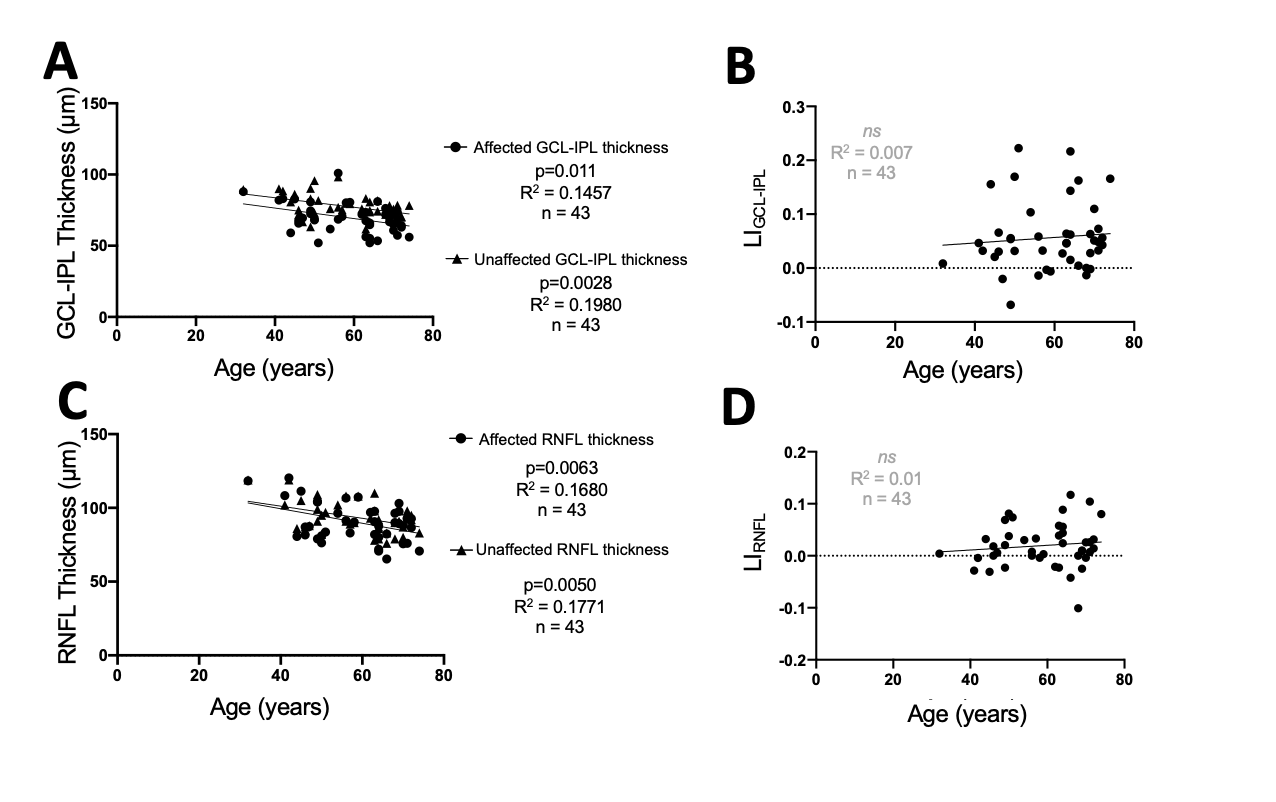

**Supplementary Figure 3.** **A:** Linear regressions of pre-training GCL-IPL thicknesses against age of participants (*affected:* *R^2^=0.1457, CI_95_(y-intercept)=74.35 to 108.1, p=0.011; unaffected: R^2^=0.1980, CI_95_(y-intercept)=84.38 to 109.8, p=0.002).* **B:** Linear regression of pre-training LI_GCL-IPL_ against age of participants (*R^2^=0.007, CI_95_(y-intercept)=-0.0863 to 0.1386, p=0.58).* **C:** Linear regressions of pre-training RNFL thicknesses against age of participants (*affected:* *R^2^=0.1680, CI_95_(y-intercept)=98.47 to 139.9, p=0.006; unaffected: R^2^=0.1771, CI_95_(y-intercept)=100.9 to 134.4, p=0.005).* **D:** Linear regression of pre-training LI_RNFL_ against age of participants (*R^2^=0.0133, CI_95_(y-intercept)=-0.0806 to 0.0665, p=0.46).*
